# Risk of Myocarditis from COVID-19 Infection in People Under Age 20: A Population-Based Analysis

**DOI:** 10.1101/2021.07.23.21260998

**Authors:** Mendel E. Singer, Ira B. Taub, David C Kaelber

## Abstract

**Background:** There have been recent reports of myocarditis (including myocarditis, pericarditis or myopericarditis) as a side-effect of mRNA-based COVID-19 vaccines, particularly in young males. Less information is available regarding the risk of myocarditis from COVID-19 infection itself. Such data would be helpful in developing a complete risk-benefit analysis for this population.

**Methods:** A de-identified, limited data set was created from the TriNetX Research Network, aggregating electronic health records from 48 mostly large U.S. Healthcare Organizations (HCOs). Inclusion criteria were a first COVID-19 diagnosis during the April 1, 2020 - March 31, 2021 time period, with an outpatient visit 1 month to 2 years before, and another 6 months to 2 years before that. Analysis was stratified by sex and age (12-17, 12-15, 16-19). Patients were excluded for any prior cardiovascular condition. Primary outcome was an encounter diagnosis of myocarditis within 90 days following the index date. Rates of COVID-19 cases and myocarditis not identified in the system were estimated and the results adjusted accordingly. Wilson score intervals were used for 95% confidence intervals due to the very low probability outcome.

**Results:** For the 12-17-year-old male cohort, 6/6,846 (0.09%) patients developed myocarditis overall, with an adjusted rate per million of 450 cases (Wilson score interval 206 - 982). For the 12-15 and 16-19 male age groups, the adjusted rates per million were 601 (257 - 1,406) and 561 (240 - 1,313).

For 12-17-year-old females, there were 3 (0.04%) cases of myocarditis of 7,361 patients. The adjusted rate was 213 (73 - 627) per million cases. For the 12-15- and 16-19-year-old female cohorts the adjusted rates per million cases were 235 (64 - 857) and 708 (359 - 1,397).

The outcomes occurred either within 5 days (40.0%) or from 19-82 days (60.0%).

**Conclusions:** Myocarditis (or pericarditis or myopericarditis) from primary COVID19 infection occurred at a rate as high as 450 per million in young males. Young males infected with the virus are up 6 times more likely to develop myocarditis as those who have received the vaccine.

## Background

Evidence has accumulated that myocarditis (used throughout, as in other studies, to include myocarditis, pericarditis or myopericarditis) is a rare side-effect of mRNA-based COVID-19 vaccines.^1-4^ A recent update from the Advisory Committee on Immunization Practices (ACIP) included an analysis that weighed the benefits of COVID-19 vaccination through reduction in infections against the harm of the vaccine. The report used data from the Vaccine Adverse Events Reporting System (VAERS) and reported rates of myocarditis following mRNA vaccination. Risk for males under 30 was about 10 times that of males age 30 or over. The highest risk group was 12-17-year-old males after the 2nd dose, with an estimated 66.7 cases per million.^4^ However, it is not known how this compares to the risk from the virus itself. Since relatively early in the pandemic, myocarditis has been recognized as a serious complication in hospitalized COVID-19 patients, including those with no prior history of cardiovascular disease, and there is histological evidence that inflammation and myocardial necrosis is present in the hearts of patients who succumbed to the disease.^5-7^ However, myocarditis is not well studied in low risk COVID-19 patients. Several recently published papers have studied myocarditis in athletes, both college and professionals, but there is limited data on risk of myocarditis in other young people.^8-10^ Although the ACIP report clearly demonstrated that the benefit of COVID-19 vaccination outweighs the risk in all age groups, a direct comparison of risk of myocarditis from disease vs. vaccination might allow for a more comprehensive risk-benefit assessment.

## Methods

A de-identified, limited data set was extracted on June 24, 2021 from the TriNetX Research Network, a federated health research network that aggregates electronic health records from 53 mostly large U.S. Healthcare Organizations (HCOs), including over 60 million people. For this study, the necessary data was available from 48 of the participating HCOs. The index date was the first COVID-19 encounter diagnosis or positive virus test, April 1, 2020 - March 31, 2021. We considered three age cohorts: 12-17 years old to match US myocarditis data following mRNA COVID-19 vaccination, and 12-15 and 16-19 years old to align with common age groupings for vaccination policy and an Israeli report of especially high risk in males age 16-19 following mRNA COVID-19 vaccination.^1,2,11^ Given the demographics of earlier reports of mRNA vaccine-associated myocarditis, our report focuses on young males, but we include data for female cohorts for comparison purposes. To ensure a reasonable diagnosis history and relationship with the HCO, patients were required to have two unrelated outpatient visits: 1 month to 2 years before the index date, and another 6 months to 2 years before that. Patients were excluded for any prior cardiovascular condition, or if they received an mRNA vaccine prior to diagnosis of myocarditis. Anyone with a diagnosis of “other specified viral infection” was excluded if they lacked a positive COVID-19 virus or antibody test. Patients with a lone COVID-19 diagnosis and a proximal (±3 days) negative COVID test but no positive test within 14 days following COVID-19 diagnosis were also excluded. Only birth year was available. Birthday was assumed to have passed if the index date was on or after July 1. The primary outcome was diagnosis of myocarditis within 90 days. Table 1 shows the ICD-10-CM diagnosis codes used. The probability of myocarditis was expected to be too low to rely on the Normal approximation to the Binomial distribution, so Wilson Score intervals were used in place of 95% confidence intervals based on the normal approximation.

**Table 1.** ICD-10-CM Diagnosis Codes.

| Condition | ICD-10-CM Diagnosis Codes |
| --- | --- |
| COVID-19 | B34.2, B97.29, J12.81, J12.82, U07.1, U07.2 |
| Myocarditis (including pericarditis, myopericarditis) | B33.22, B33.23, I30.1, I40, I41, I51.4 |
| Cardiovascular condition | B33.22, B33.23, I00-I99, R00, R01.1, R01.2, R03 |

While cases of myocarditis are expected to result in interaction with the health care system, there will be many missed COVID-19 cases that will not be detected in the HCO’s electronic health records. We sought to estimate the proportion of missed COVID-19 cases. For each cohort, we queried the TriNetX Live Research Network to find people with matching demographics using similar engagement with the HCO: outpatient visit in the system in the two years ending March 31, 2021 (our study’s ending date for index COVID-19 diagnosis or positive virus test), as well as another outpatient visit six months to two years before that. Of these people, we queried to find out how many of them had a COVID-19 diagnosis or positive virus test in our study period, April 1, 2020 - March 31, 2021. This proportion was compared to the proportion of the population that had COVID-19 during that period. In the United States, there is no national data on infection rate by age. According to a systematic review and the working assumption of the Centers for Disease Control and Prevention, children may have infection rates similar to adults, with younger people having more mild or asymptomatic cases.^12,13^ Accordingly, we used the estimated 9.2% population infection rate for April 2020 - March 2021.^14^ The estimated proportion of COVID-19 cases for 12-17 year old males was 2.5%. We then multiplied the denominator of COVID-19 cases in our study by 3.7 (9.2 / 2.5) to arrive at an adjusted number of COVID-19 cases. Similar calculations were done for the other cohorts.

The missed COVID-19 cases can be broken down into three categories: not tested and no physician contact; tested outside the TriNetX system but no physician contact; and tested and received care outside the TriNetX system. We assumed this last group would have clinical courses similar to those followed up in the TriNetX database. All three groups of missed cases were expected to be substantial in size and in the absence of data to refine the estimate, we assumed all three groups would be of equal size. Based on these assumptions, we adjusted the number of myocarditis cases to reflect the missed cases arising from patients tested and followed up outside of TriNetX.

## Results

For the 12-17-year-old male cohort, 6,846 patients met the study criteria (Table 2). There were 6 (0.09%) cases of myocarditis overall, corresponding to a rate of 1 case of myocarditis per 1,141 COVID-19 patients, or 876 cases per million patients (Wilson Score interval: 402 - 1,911). After adjusting for missed cases of COVID-19 and myocarditis, the adjusted cases per million was 450 (206 - 982).

**Table 2.** Rates of Myocarditis in a Sample of COVID-19 Infected Patients.

|  | **N**<br>**(COVID-19)** | **Myocarditis**<br>**N (%)** | **Rate per Million**<br>**(Wilson Score Interval)**<br>**# (Confidence Interval)** | **Adjusted Rates per**<br>**Million**<br>**# (Confidence**<br>**Interval)** |
| --- | --- | --- | --- | --- |
| **Age 12-17 Males** | 6,846 | 6 (0.09%) | 876 (402 - 1,911) | 450 (206 - 982) |
| **Age 12-15 Males** | 4,114 | 5 (0.12%) | 1,215 (519 - 2,842) | 601 (257 - 1,406) |
| **Age 16-19 Males** | 5,097 | 5 (0.10%) | 981 (419 - 2,294) | 561 (240 - 1,313) |
| **Age 12-17 Females** | 7,361 | 3 (0.04%) | 408 (139 - 1,198) | 213 (73 - 627) |
| **Age 12-15 Females** | 4,280 | 2 (0.05%) | 467 (128 - 1,702) | 235 (64 - 857) |
| **Age 16-19 Females** | 6,687 | 8 (0.12%) | 1,196 (606 - 2,359) | 708 (359 - 1,397) |
| **Age 12-17, All** | 14,207 | 9 (0.06%) | 633 (333 - 1,204) | 328 (173 - 624) |
| **Age 12-15, All** | 8,394 | 7 (0.08%) | 834 (404 - 1,721) | 416 (202 - 858) |
| **Age 16-19, All** | 11,784 | 13 (0.11%) | 1,103 (645 - 1,887) | 643 (376 - 1,100) |

For the 12-15 and 16-19 male age groups, the number of myocarditis cases were 5 of 4,114 (0.12%) and 5 of 5,097 (0.10%), and the adjusted cases per million were 601 (257 - 1,406) and 561 (240 - 1,313).

For 12-17-year-old females the adjusted rate of myocarditis was 213 (73 - 627) per million cases. For the 12-15- and 16-19-year-old female cohorts the adjusted rates per million cases were 235 (64 - 857) and 708 (359 - 1,397).

When males and females were combined the adjusted rates per million cases for age 12-17, 12-15 and 16-19 were 328 (173 - 624), 416 (202 - 858) and 643 (376 - 1,100), respectively.

8/20 (40.0%) cases of myocarditis were diagnosed within 5 days of the index date, while the other 12/20 (60.0%) cases were diagnosed 19-82 days after the index date. Two patients were hospitalized, one and three days after the index date. There were no reported deaths.

## Discussion

Based on existing reports, myocarditis after RNA COVID-19 vaccination occurs largely after the second dose. The highest risk subgroup is 12-17 year old males, with 66.7 cases per million second doses and 9.8 per million first doses for a combined total of 76.5 cases per million vaccine recipients.^1,2^, Our results suggest that, even for this high-risk subgroup, the risk of myocarditis from COVID-19 infection is about 5.9 times as great, at a rate of 450 cases per million. Based on the background rate of myocarditis in this population, the expected rate in the absence of COVID-19 for 90 days would be less than 0.1.^15^ For 12-17 year old females, myocarditis following mRNA COVID-19 vaccination was 1.1 and 9.1 per million following the first and second doses, for a total of 10.2 per million getting vaccinated.^4^ Risk of myocarditis from COVID-19 infection was nearly 21 times that rate, with an adjusted rate of 213 cases per million. For both males and females, risk of myocarditis from COVID-19 infection was higher in the 16-19 year-old than the corresponding 12-17 cohort.

Time from COVID-19 diagnosis or positive virus test to myocarditis was split into two distinct groups: 8/20 (40.0%) within 5 days and the rest at 19-82 days following index COVID-19 diagnosis or positive virus test. This may represent a combination of acute COVID-19 myocarditis and post-COVID-19 myocarditis. However, some of the delayed diagnoses may represent patients hospitalized outside the TriNetX system who later followed up with their primary care provider in the HCO. Two patients had no COVID-19 diagnosis, but diagnosis of myocarditis at 62 and 82 days after positive COVID-19 virus test. One had their first diagnosis of both COVID-19 and myocarditis 58 days after positive virus test. Three others had their first COVID-19 diagnosis in the system at 5, 13 and 32 days after positive virus test and myocarditis diagnosed at days 20, 63 and 40, respectively. It is likely that some of these 6 cases represent people who were hospitalized at a facility not in the TriNetX database, making the true hospitalization rate perhaps several times higher than the 2/20 (10.0%) in the database, though any hospitalizations are in the context of other COVID-19 symptoms and complications. Whatever the true hospitalization rate was, it was considerably lower than that reported in the VAERS, where more than three-fourths of reported cases of myocarditis were hospitalized.^16^. Cases have been described as generally mild.^2,17,18^ Admission rates may settle at a much lower rate over time as the natural history becomes better understood.

Several studies have identified post-COVID19 myocarditis in collegiate and professional athletes at rates of 0.6 - 2.3%.^8-10^ However, there are comprehensive testing protocols for high level collegiate and professional athletes that are not applicable to the general population, and these may over-estimate the rate of clinically significant disease. One study of college athletes at Big-Ten schools reported that if a published diagnostic strategy based on cardiac symptoms had been employed, just 0.31% would have been diagnosed.^9,19^ In this study, rates were about 0.1% before downward adjustments for missed COVID-19 cases. This lower rate is likely due to some combination of missed cases for reasons explained above, and a somewhat younger population.

Reliance on diagnosis codes for myocarditis might underestimate or overestimate cases. More than 1 in 5 COVID-19 cases were based only on diagnosis codes. This may be due to presumed diagnosis based on a combination of symptoms and family exposure. It may also be due in part to testing that takes place outside the HCO and is not in the HCO’s EHR, e.g. pharmacies, community testing sites, health departments. Myocarditis cases may also be missed due to insufficient follow-up for COVID-19 cases at the end of the study period.

Another limitation is the approach taken to account for missed cases of COVID-19. We assumed that infection rates are similar for 12-19-year-olds and the overall population, and that one-third of the extra COVID-19 cases not detected in the database were tested and seen by physicians with similar rates of myocarditis. There is no currently available data to support precise estimates. However, assuming no additional cases of myocarditis from any of the missed COVID-19 cases, rates of myocarditis in 12-17 year old males would still be nearly three times as great from COVID-19 infection than from the vaccine.

With intense media and social media focus on COVID-19 vaccine side-effects, it is important to quantify and communicate to the public the risks of COVID-19 infection in young people. The ACIP report projected that mRNA vaccination in 12-17-year-old males would result in 215 fewer hospitalizations and 71 fewer intensive care unit stays. Benefits of the vaccine outweighed the risk of myocarditis from vaccination in all age groups, 12 years-old and up. Our results suggest that the risk of myocarditis from COVID-19 infection itself exceeds the known risk from vaccination by a considerable margin. In light of more infectious variants, the new school year nearing and many colleges now requiring COVID-19 vaccination (either for all students or just those living on campus), these results are especially timely. Whether considering all the risks and benefits of COVID-19 vaccination or just myocarditis, vaccination appears to be the safer choice for 12-19-year-old males and females.

## Data Availability

The data set is licensed and is not publicly available.

## Acknowledgement

This publication was made possible by the Clinical and Translational Science Collaborative of Cleveland, UL1TR0002548 from the National Center for Advancing Translational Sciences (NCATS) component of the National Institutes of Health and NIH roadmap for Medical Research. Its contents are solely the responsibility of the authors and do not necessarily represent the official views of the NIH.

